# The Right to Sexual and Reproductive Health among Migrant Workers in Taiwan: Stakeholder Perspectives through an AAAQ Analysis

**DOI:** 10.64898/2026.08.26.26361458

**Authors:** Patrick Tang, Michelle Wei-Hsuan Lu, Kai-Tsun Yeung, Bruce Jiajie Guo, Ko-Fei Natalie Wei

**Affiliations:** Harvard University, Department of Global Health and Population, Boston, USA; National Taiwan University, College of Medicine, Taipei, Taiwan; University of Michigan, Ann Arbor, USA; Emory University, Atlanta, USA; TES, Taipei, Taiwan

**Keywords:** Migrant workers, Sexual and reproductive health and rights, AAAQ framework, Health equity, Taiwan

## Abstract

**Background:** Global labor migration from LMIC to higher-income destinations has expanded rapidly, placing increasing pressure on destination-country health. Existing research on cross-border migrant workers has focused largely on occupational health, general healthcare utilization, and disease-specific risks, while there is considerably less evidence on their sexual and reproductive health. This study contributes to this understudied field by examining the policy and health-system factors that shape the sexual and reproductive health services for migrant workers in Taiwan.

**Methods:** A qualitative study was conducted in Taiwan between November 2025 and August 2026. 22 stakeholders were purposively recruited from academia, healthcare, nongovernmental organizations, government, labor brokerage, and employers. Data were collected through semi-structured interviews and small focus groups. Interviews were conducted in Mandarin Chinese, transcribed verbatim, and translated into English. Data were analyzed using framework analysis combining deductive coding based on the AAAQ framework with inductive coding of implementation and contextual themes.

**Results:** Gaps were identified across all four AAAQ dimensions. Participants described limited migrant- responsive SRH programming; physical, financial, administrative, social, and information barriers; shortcomings in linguistic and cultural responsiveness; and weaknesses in interpretation, coordination, and continuity of care, despite generally favorable views of Taiwan’s clinical quality.

**Conclusions:** Our findings show that broad insurance coverage and strong clinical capacity do not by themselves ensure the realization of migrant workers’ SRHR. In Taiwan, rights were mediated through labor brokerage, gendered live-in work arrangements, and fragmented governance across health, labor, immigration, and social-welfare systems. Improving migrant SRHR therefore requires stronger implementation of existing protections, reduced dependence on informal intermediaries, and more integrated institutional responsibility for cross-sector migrant health needs.

## 1. Introduction

Migrant workers are among the groups most vulnerable to gaps between the formal recognition and practical realization of the right to health. Despite growing international commitments to universal health coverage and health equity, migrants continue to experience disproportionate barriers to accessing healthcare due to language differences, legal and administrative restrictions, discrimination, financial constraints, and mobility. [1–3] These constraints are especially pronounced among low-skilled, employment-based migrant workers, whose access to care is often shaped by labor dependency, employer control, and exclusion from routine health protections. [4] As a result, these barriers often translate into delayed care, lower utilization of preventive services, poorer health outcomes, and widening health inequities, even in countries with otherwise well-developed health systems. [1, 5]

Sexual and reproductive health (SRH) presents additional implementation challenges beyond those encountered in general healthcare. Unlike many other health services, SRH is closely intertwined with stigma, privacy, gender norms, cultural and religious beliefs, and concerns regarding confidentiality and discrimination.[6] Consequently, access to SRH information, prevention, testing, treatment, and care is often influenced not only by the availability of services but also by whether individuals feel safe, respected, and empowered to seek care without fear of social, legal, or employment-related consequences. [7, 8]

Thus, when seeking SRH services, migrant workers must navigate two intersecting systems of structural barriers. Consistent with theories of intersectionality and compounded vulnerability, these barriers are not merely additive; rather, they interact to shape experiences of exclusion that are qualitatively different from those associated with migration or SRH alone. [9–11] This makes migrant workers’ SRH a critical site for examining how rights-based health commitments break down at the point of implementation.

Taiwan offers an ideal setting to study this unique implementation challenge. Over the past two decades, Taiwan has progressively strengthened SRH protections through legal reforms, expansion of HIV and STI prevention programs, and broader efforts to promote gender equality and sexual rights. [12] At the same time, Taiwan relies on a large migrant workforce of more than 800,000 workers employed primarily in manufacturing, construction, agriculture, fisheries, domestic work, and long-term care. [13]

Nevertheless, despite their increasing, structural importance to Taiwan’s economy and welfare system, migrant workers’ SRH remains understudied. Existing studies of migrant workers have primarily examined occupational safety, mental health, and general healthcare utilization, while Taiwan’s SRH literature has largely focused on HIV epidemiology, behavioral risk, and specific priority populations such as men who have sex with men. [14–21] As a result, relatively little is known about migrant workers’ access to SRH information, prevention, testing, treatment, and care. Additionally, no published study to our best knowledge has examined this intersection through a rights-based lens, leaving unresolved how legal protections, health policies, and service delivery arrangements shape the practical realization of sexual and reproductive health and rights (SRHR) for migrant workers.

To address this gap, this study adopts the Availability, Accessibility, Acceptability, and Quality (AAAQ) framework articulated by the United Nations Committee on Economic, Social and Cultural Rights. [22] Widely applied in rights-based evaluations of health systems, the AAAQ framework operationalizes the right to health by examining whether services are sufficiently available, physically and economically accessible, culturally acceptable, and of adequate quality. [22] Using Taiwan as a case study, we conducted a qualitative policy analysis based on semi-structured interviews with key stakeholders responsible for the governance, financing, implementation, and delivery of migrant workers’ SRH services, including representatives from government agencies, healthcare providers, legislators, employers, labor brokers, and civil society organizations. By examining institutional perspectives across the policy implementation process, this study seeks to identify where implementation gaps arise and how they shape the realization of SRHR for migrant workers.

This study makes three contributions to the literature. First, it addresses an important empirical gap by examining the largely overlooked intersection of migrant workers and SRH in Taiwan. Second, it advances a rights-based perspective on migrant SRH by moving beyond conventional epidemiological and behavioral approaches to examine how legal protections, governance arrangements, and service delivery interact to facilitate or constrain the realization of SRHR. Finally, by applying the AAAQ framework to the perspectives of policy and implementation stakeholders, this study provides a systems-level understanding of where implementation gaps emerge and identifies actionable opportunities to strengthen the governance of migrant SRHR in both Taiwan and other similar contexts.

## 2. Methods

This study was conducted between November 2025 and August 2026 to examine the governance of SRHR for migrant workers in Taiwan. Purposive sampling was used to recruit stakeholders occupying different positions within the governance and implementation ecosystem of migrant SRH, including academia, healthcare providers, NGOs, government agencies, labor brokers, and employers. A total of 22 participants were selected based on their direct involvement in developing, implementing, delivering, financing, or facilitating SRH services and policies for migrant workers. To capture both organizational and cross-sector perspectives, data were collected through a combination of individual semi-structured interviews and small focus groups. (*See Table 1*)

**Table 1:** Participant Information.

| Stakeholder category | Participant code | Organization type / role | Data collection method |
| --- | --- | --- | --- |
| Academia | AC01 | University researcher involved in migrant health implementation projects | Individual interview |
| Academia | AC02 | University researcher involved in HIV policy research | Individual interview |
| Healthcare | HC01 | Migrant-facing healthcare provider | Focus group discussion 1 |
| Healthcare | HC02 | Migrant-facing healthcare provider | Focus group discussion 1 |
| Healthcare | HC03 | STI/HIV clinical service provider | Individual interview |
| Healthcare | HC04 | STI/HIV clinical service provider | Individual interview |
| NGO | NGO01 | HIV/AIDS service organization | Individual interview |
| NGO | NGO02 | HIV/AIDS service organization | Individual interview |
| NGO | NGO03 | HIV/AIDS service organization | Individual interview |
| **NGO** | NGO04 | Migrant worker support organization | Focus group discussion 2 |
| **NGO** | NGO05 | Migrant worker support organization | Focus group discussion 2 |
| **NGO** | NGO06 | Migrant worker support organization | Focus group discussion 2 |
| **NGO** | NGO07 | Migrant worker support organization | Individual interview |
| **NGO** | NGO08 | HIV/AIDS service organization | Individual interview |
| **Government** | GOV01 | Legislative stakeholder | Individual interview |
| **Government** | GOV02 | Public health authority | Individual interview |
| **Government** | GOV03 | Local labor administration stakeholder | Individual interview |
| **Government** | GOV04 | Immigration administration stakeholder (former staff) | Individual interview |
| **Labor broker** | BK01 | Employment brokerage agency | Focus group discussion 3 |
| **Labor broker** | BK02 | Employment brokerage agency | Focus group discussion 3 |
| **Employer** | EM01 | Employer of migrant workers | Individual interview |
| **Employer** | EM02 | Employer of migrant workers | Individual interview |

Interviews were conducted in Mandarin Chinese either in person or via Zoom and generally lasted 30–60 minutes. The semi-structured interview guide was developed using the Availability, Accessibility, Acceptability, and Quality (AAAQ) framework. All participants provided verbal informed consent before participation. Interviews were audio-recorded, transcribed verbatim, translated into English, and de-identified before analysis. Ethical approval was obtained from Harvard Longwood Campus Institutional Revision Board (IRB25- 1054) under exempt review.

Data were analyzed using framework analysis, combining deductive and inductive coding. An initial coding framework was developed a priori based on the AAAQ framework and operationalized into a hierarchical codebook comprising the four primary domains of availability, accessibility, acceptability, and quality. Accessibility was further disaggregated into five subdomains—physical, financial, administrative, social, and information accessibility—to capture the multidimensional barriers influencing migrant workers’ access to SRH services. During analysis, inductive codes were iteratively incorporated to capture implementation challenges and contextual factors that were not fully represented within the predefined framework. Four researchers independently coded the transcripts and resolved discrepancies through discussion, with the first author serving as the final arbiter of coding decisions. Coding and data collection proceeded iteratively until thematic saturation was achieved, with no substantively new implementation themes emerging from subsequent interviews.

## 3. Results

Across the AAAQ framework, participants identified gaps in the availability, accessibility, acceptability, and quality of SRH services for migrant workers. These gaps reflected not only limitations in service provision, but also barriers arising from employment conditions, language and cultural differences, administrative complexity, and fragmented coordination across health and labor systems. The following sections present findings within each AAAQ domain. (*See Table 2*)

**Table 2:** Selected Quotes for AAAQ Analysis.

| AAAQ | Subdomain | Participant | Stakeholder | Context | Selected quote |
| --- | --- | --- | --- | --- | --- |
| **Availability** | Type of SRH services | AC01 | Academic researcher | Sexual health education and contraceptive information channels for migrant workers | "Broader sex education, or to say, the provision of contraception information channels, as far as I know, is also relatively less, as if this is not an aspect that the Ministry of Labor would lean toward emphasizing." |
| **Availability** | Type of SRH services | AC01 | Academic researcher | Contraceptive methods available to Indonesian female migrant workers in Taiwan | "In Taiwan's medical institutions, injectable contraception is also not that easy to obtain... upper-arm implant contraceptive device in Taiwan it is also not common." |
| **Availability** | Quantity of SRH services | NGO01 | HIV/AIDS NGO | HIV-related services for foreign nationals and migrant workers | "There are actually very few NGOs that can provide HIV-related services—mainly us and (another NGO)." |
| **Availability** | Quantity of SRH services | NGO03 | HIV/AIDS NGO | Shelter and practical support services for migrant workers living with HIV | "If all the beds are occupied, then we have to look for other partners." |
| **Accessibility** | Physical accessibility | HC04 | Healthcare provider | Live-in domestic caregivers seeking healthcare | "For caregivers it is much harder, because they work one-on-one and have to be beside the patient almost all the time. It is very difficult for them to take leave." |
| **Accessibility** | Physical accessibility | EM01 | Employer | Migrant caregivers attending prenatal appointments | "If a migrant worker needs to go for a prenatal checkup, I have to arrange substitute long-term care first... if long-term care has no availability... the worker cannot postpone the appointment." |
| **Accessibility** | Physical accessibility | NGO04–06 | Migrant support NGO | Migrant workers accessing healthcare outside metropolitan areas | "Unless migrant workers are in major cities, it is often difficult for them to find legal and accessible healthcare services." |
| **Accessibility** | Financial accessibility | NGO03 | HIV/AIDS NGO | HIV treatment costs for migrant workers living with HIV | "The monthly medication cost is over NT\$10,000, which many of them simply cannot afford." |
| **Accessibility** | Financial accessibility | BK02 | Labor broker | Uptake of self-paid preventive healthcare among migrant workers | "If they have to pay, they will not get it, but if it is free, they definitely will." |
| **Accessibility** | Financial accessibility | AC02 | Academic researcher | Income loss among pregnant migrant workers receiving shelter support | "The mom will be very hard-pressed because she has no work... from waiting to give birth to the first two months of the child, this is completely zero income." |
| **Accessibility** | Administrative accessibility | GOV03 | Local labor bureau staff | Coordination of migrant workers' healthcare, employment, and social protection issues | "There are too many separate regulations, authority is fragmented." |
| **Accessibility** | Administrative accessibility | HC01–02 | Healthcare provider | Implementation of migrant worker health and labor protections | "The main problem with policy is that implementation gets stuck at the employer level." |
| **Accessibility** | Administrative accessibility | NGO03 | HIV/AIDS NGO | HIV treatment access and immigration-related barriers among foreign nationals | "The visa and immigration issues then become even more complicated. It depends on their status and their particular circumstances." |
| **Accessibility** | Social accessibility | AC02 | Academic researcher | Pregnancy decision-making among migrant workers | "The most extreme example I encountered... was the employer not only disagreed, he also requested the migrant worker to go have an abortion, and then requested the broker to take the migrant worker to have an abortion." |
| **Accessibility** | Social accessibility | NGO02 | Migrant support NGO | Pregnant migrant workers facing employer/broker pressure | "Brokers or employers... will try every possible way to get pregnant migrant mothers to 'voluntarily' sign papers, terminate their contracts, and return home... There is often a mix of persuasion and deception that pushes migrant workers to sign and leave." |
| **Accessibility** | Social accessibility | NGO01 | HIV/AIDS NGO | Foreign nationals/migrant workers living with HIV | "Many people are reluctant [to be reported] because they worry that this status will follow them permanently. They also worry about practical consequences—such as their job [and] their future prospects." |
| **Accessibility** | Information accessibility | AC02 | Academic researcher | Communication between migrant workers and healthcare professionals regarding medication and health advice | "I as the person seeking service, have difficulty in my expression, and then... this medical professional worker... also has difficulty in giving professional knowledge, or the medication knowledge given by the pharmacist." |
| **Accessibility** | Information accessibility | AC01 | Academic researcher | Dissemination of contraceptive information among Indonesian migrant workers | "Especially Indonesian migrant workers, her channel of disseminating information actually is just TikTok... if we have information we want to pass to them, but we don't go through... the channels they usually receive information through, then it is not quite possible to transmit to her." |
| **Accessibility** | Information accessibility | GOV01 | Legislative Staff | Migrant workers navigating Taiwan's healthcare system | "They may not know when to go to a clinic and when to go to a hospital... what documents to bring, how to register, what co-payments are, or which services may not be covered." |
| **Acceptability** | Linguistic respect & communication | HC01–02 | Healthcare provider | Migrant workers receiving healthcare services | "Some may even feel that speaking English to them is a kind of class-based mockery... an elite language they are not comfortable with. That is why using Tagalog can be more respectful and more comfortable." |
| **Acceptability** | Cultural relevance of health education | NGO02 | Migrant support NGO | Migrant workers receiving sexual health education | "For people from other countries, it is not enough simply to take the same version and translate it into different languages." |
| **Acceptability** | Compatibility of contraceptive practices | AC01 | Academic researcher | Indonesian female migrant workers accessing contraceptive services in Taiwan | "Migrant workers are not that they don't want to contracept... after she came to Taiwan there is a gap, and then there is also a process of transition, and then plus language, culture, work style." |
| **Quality** | Clinical quality and patient experience | HC01–02 | Healthcare provider | Migrant workers' experiences with Taiwan's healthcare system | "Migrant workers' highest satisfaction with Taiwan is with health care... they have the best impression of Taiwan's medical care." |
| **Quality** | Communication quality | HC03 | Healthcare provider | Healthcare communication with migrant workers | "Medical interpretation is also extremely lacking." |
| **Quality** | Continuity & coordination of care | NGO03 | HIV/AIDS NGO | HIV treatment continuity among migrant workers | "If there were delays [in importing medication], then their treatment regimen might be interrupted, leading to drug resistance and affect their treatment outcomes." |

### 3.1 Availability

Within the AAAQ framework, availability concerns whether health facilities, services, programs, personnel, medicines, information, and related resources exist in sufficient type and quantity to meet the health needs of a population. [23] Participants identified limitations both in the range of SRH services available to migrant workers in Taiwan and in the number and capacity of organizations providing them.

Regarding the types of services available, participants described gaps in government provision. Broader sexuality education and channels for obtaining contraceptive information were perceived to receive limited attention within labor-related programming. As one academic participant explained, the provision of contraceptive information was “relatively less,” and did not appear to be an area that the Ministry of Labor would “lean toward emphasizing” (AC01). The same participant noted that injectable contraceptives were “not that easy to obtain” in Taiwanese medical institutions and that upper-arm contraceptive implants were uncommon (AC01). Local health bureau outreach for HIV and sexually transmitted infection screening was also reported to focus on new immigrants and foreign spouses while overlooking foreign workers (BK02). One NGO representative identified a further gap in programming, observing that whether migrant workers’ sexual needs could be met, and how they could be met safely, were issues “we (as a society) simply do not discuss” (NGO03).

The services identified in the data were provided predominantly through nongovernmental and shelter-based initiatives. These included preventive HIV education and practical care for migrant workers living with HIV, shelter-based health education, temporary accommodation, basic material assistance, maternal and child health consultation, and residential support for migrant workers with broader health and social needs (HC01–02; NGO02; NGO03; NGO04– 06). For example, one shelter offered daily classes covering health education and infectious diseases and maintained designated residential space for mothers (HC01–02). One NGO operated a maternal and child health consultation center in Taoyuan specifically for foreign women and children, alongside two shelters serving migrant workers with diverse needs (NGO04–06). Another NGO divided its migrant-worker services between preventive education and practical care for those living with HIV (NGO03). These services were linked to particular organizations rather than described as components of a broadly available government program (HC01–02; NGO02; NGO03; NGO04–06). Reflecting this residual role, one NGO characterized its assistance as practical “last-mile help” provided “at the very end of the chain” (NGO08).

Some relevant resources were available through healthcare institutions or cross-organizational collaboration. For example, one hospital offered free interpretation services, but a healthcare participant reported that staff “did not really know which unit to apply to or how to apply” (HC04). The same participant described the collaborative production of a mental-health handbook in Indonesian, Filipino, and Vietnamese (HC04). Another healthcare participant reported involvement in migrant-worker free clinics, while emphasizing that much of the bridging work continued to rely on NGOs and church groups because “no official unit is devoted to doing that bridging work” (HC03). These accounts point to discrete institutional and collaborative initiatives rather than a dedicated government system of sexual health provision for migrant workers (HC03; HC04).

Regarding quantity and capacity, participants emphasized the small number of specialized providers. One NGO representative stated that “there are actually very few NGOs that can provide HIV-related services,” identifying their organization and one other NGO as the principal providers in this area (NGO01). The organization assisted approximately 100–120 individuals annually, but the participant reported that “there are no dedicated programs specifically supporting foreign nationals or migrant workers” (NGO01). Material capacity was also finite. One NGO had to seek assistance from partner organizations “if all the beds are occupied” (NGO03). Another reported that direct linguistic support was limited to English, French, and Tibetan; beyond those languages, “we really cannot do much—unless the person has their own interpreter” (NGO07).

Participants also described limited government infrastructure supporting migrant workers’ sexual health. A government participant argued that, although individual ministries could point to existing regulations or healthcare services, employers and brokers were left with frontline responsibility when concrete needs arose (GOV01). The participant therefore called for the government to build public-service infrastructure encompassing independent medical advice, interpretation, case management, and cross-ministerial coordination (GOV01).

### 3.2 Accessibility

The AAAQ right-to-health framework distinguishes five interrelated dimensions of accessibility: physical, financial, administrative, social, and information. [23]

#### 3.2.1 Physical Accessibility

Physical accessibility determines whether SRH services can be reached in practice, including whether their location, operating hours, transportation requirements, and time demands are compatible with migrant workers’ living and working conditions. Participants described work schedules and restrictions on leaving the workplace as major barriers to reaching care. Live-in caregivers were reported to have little time away from the household and to require broker notification before attending medical appointments (HC01–02). Because caregivers often had to remain beside care recipients, taking leave could be particularly difficult; as one healthcare participant stated, “what they lack most now is time” to seek care or otherwise attend to their own health needs (HC04). Migrant workers’ days off were also reported not to align with hospital or clinic operating hours (NGO04–06).

Access to prenatal and follow-up care could depend on whether employers were able to arrange substitute care or authorize leave. One participant explained that a caregiver could not attend a prenatal appointment unless replacement long-term care was first secured, yet substitute services were not always available (BK02; EM01). Participants similarly questioned how workers requiring appointments every three to six months could repeatedly obtain leave from their employers (NGO07; NGO08).

Geographic location and navigation created additional barriers. Migrant workers outside major cities were described as having difficulty locating lawful and reachable healthcare services, particularly those working in remote fishing ports (HC03; NGO04–06). Even after reaching an urban hospital, workers could face difficulties identifying the correct transit exit, building, or department (HC04). These barriers were compounded when workers were unable to read directions or were reluctant to attend hospital alone, creating additional transportation, accompaniment, and staffing needs for NGOs (HC01–02; NGO07).

#### 3.2.2 Financial Accessibility

Despite the broad affordability of healthcare under Taiwan’s National Health Insurance (NHI) system, participants described important gaps in financial protection for migrant workers, particularly for services excluded from coverage and during periods of interrupted employment. These barriers included direct medical expenses, uncertainty regarding insurance coverage, and income loss associated with pregnancy or illness.

The most substantial direct costs related to HIV treatment. In Taiwan, NHI subsidies for HIV treatment are generally available to foreign nationals only after they have been enrolled in the NHI system for more than two years. During this initial two-year period, migrant workers living with HIV may therefore face substantial out-of-pocket treatment costs (NGO01; NGO07; NGO08). Participants estimated monthly medication costs at more than NT$10,000, with some citing amounts between NT$13,000 and NT$20,000 (NGO01; NGO02; NGO03). As one NGO participant explained, “the monthly medication cost is over NT$10,000, which many of them simply cannot afford” (NGO03). Although since 2024, the government has offered a support program that provide antiretroviral medication to some foreign nationals during this coverage gap, participants described the subsidies as “difficult to get” (HC04; BK02; NGO01-03; NGO07; NGO08). NGOs therefore continued to rely on charitable donations and other ad hoc assistance for workers who could not obtain sufficient support through the program (NGO01- 03; NGO07; NGO08).

Costs also influenced the uptake of preventive and reproductive health services. Several participant observed that migrant workers would accept free services but often forgo self-paid interventions, including influenza vaccination (NGO08; BK02; EM01-02). Migrant women were similarly described as needing to navigate unfamiliar contraceptive options after arrival in Taiwan (AC01). An academic participant noted that workers were sometimes unclear about the cost of intrauterine devices and whether these methods were covered by NHI (AC01).

Financial vulnerability extended beyond medical fees. Recruitment debt, continuing broker payments, and remittance obligations reduced the resources available for healthcare (HC01– 02; HC03). For pregnant workers entering shelters, the more immediate burden was often loss of income rather than medical expenditure: one participant noted that, from the period before childbirth through the first two postpartum months, women could have “completely zero income” (AC02). Although participants acknowledged that NHI substantially reduced healthcare costs, coverage gaps and employment-related income loss left some migrant workers exposed to expenses disproportionate to their earnings (AC02; BK01; GOV02).

#### 3.2.3 Administrative Accessibility

Administrative accessibility to SRH services was constrained by fragmented responsibility across labor, health, immigration, and insurance systems. Participants described cases in which no authority assumed responsibility for guiding workers through the full process of obtaining care, leave, documentation, and continued employment (GOV03; NGO01; NGO03). As one government participant explained, “there are too many separate regulations” and “authority is fragmented,” with labor authorities sometimes treating a case as a medical matter and brokers providing inconsistent answers (GOV03). The practical connection between formal policies and services therefore often depended on NGOs rather than an established interagency pathway (HC03).

A second theme was the dependence of formal protections on employer cooperation. Pregnancy alone did not lawfully justify dismissal, yet contracts could still be terminated through purported mutual agreement, and changing employers after pregnancy was described as “basically not feasible” (AC02; BK01). Participants further noted that leave, continued employment, and return-to-work arrangements were often controlled at the workplace level rather than determined solely by medical need (HC01–02; HC04). As one healthcare participant summarized, “the main problem with policy is that implementation gets stuck at the employer level” (HC01–02). This dependence was especially consequential for domestic caregivers, who could not seek care or take leave without disclosing their circumstances to employers or brokers (HC04).

Administrative access was also shaped by legal and insurance status. HIV treatment required formal diagnosis and notification, while subsequent access to publicly financed treatment depended on continued NHI eligibility and lawful residence (NGO01; NGO03; NGO07; NGO08). Loss of employment could precipitate the expiry of residence status, and undocumented status restricted access to direct financial assistance and formal placement services (NGO02; NGO03; NGO08).

#### 3.2.4 Social Accessibility

Social accessibility concerns whether healthcare can be sought and used without stigma, discrimination, coercion, or exclusion arising from social and employment relationships. Three interrelated barriers emerged: reproductive coercion, anticipated consequences of HIV disclosure, and employer-dependent social isolation.

Pregnancy-related decisions were shaped by pressure from employers and brokers. One participant described an employer requesting that a migrant worker obtain an abortion and instructing the broker to “take care of the business” (AC02). Another reported that employers and brokers used “a mix of persuasion and deception” to induce pregnant workers to sign contract-termination documents and return home (NGO02). Fear of job loss or repatriation encouraged concealment, with one participant noting that a worker might be “afraid the employer will find out and not want her” (BK01).

HIV-related stigma similarly discouraged disclosure and engagement with care. Workers were reported to fear that an HIV diagnosis would reach employers or brokers and damage their employment and future prospects (NGO01; NGO03). Although Taiwanese law prohibits dismissal solely on the basis of HIV status, one participant stated that employers often “terminate the contract or send (foreign care workers) away” after learning of a diagnosis (NGO08). The same participant described public narratives portraying migrant workers living with HIV as threats who might “infect Taiwanese people” (NGO08).

These pressures were intensified by employer dependence. Live-in caregivers were described as residing with employers “24 hours a day,” increasing exposure to control and abuse (AC01). Participants also identified sexual harassment, workplace abuse, racial prejudice, and an oppressive social environment for Southeast Asian migrant workers (HC01–02; HC03; NGO02).

#### 3.2.5 Information Accessibility

Information accessibility concerns whether people can seek, receive, and understand health information in forms and languages that support informed decision-making. [23] Participants described barriers arising from language, ineffective dissemination channels, and limited practical guidance on navigating Taiwan’s healthcare system.

Language barriers affected both written materials and clinical encounters. Hospital guidance was frequently available only in Chinese, while bilingual materials explaining procedures and required actions were described as insufficient (HC01–02; NGO04–06). Communication difficulties also operated in both directions: migrant workers experienced difficulty expressing their needs, while clinicians and pharmacists struggled to convey professional advice and medication instructions accurately (AC02). One participant emphasized that written guidance alone did not resolve this problem, because “when they face each other face-to-face, actually there still is a gap” (AC02)

Official dissemination was also poorly aligned with the channels and formats workers used. Government websites generally provided information only in Chinese and English, while translated policy materials were described as “dense and hard to understand” and not designed from users’ perspectives (HC03). One participant explained that many Indonesian migrant workers obtained information primarily through TikTok and that messages disseminated outside these established channels were “not quite possible to transmit” to them (AC01). Pregnancy-related guidance likewise circulated through TikTok and Facebook rather than direct government promotion (BK01).

Participants also described limited practical knowledge of healthcare navigation and SRH services. Workers lacked information about registration, required documents, co-payments, insurance coverage, symptoms warranting testing, and where to seek assistance (GOV04; NGO01; NGO02). Information gaps also affected contraceptive use, including awareness of intrauterine devices and the correct use of emergency contraception (AC01; BK01). NGOs partly addressed these gaps through simplified multilingual booklets and explanations of government HIV-support measures, although conflicting online and pre-departure information continued to create uncertainty (NGO03; NGO08).

### 3.3 Acceptability

Acceptability depended on whether SRH communication and services were linguistically respectful, culturally relevant, and compatible with workers’ prior experiences. A healthcare participant cautioned against assuming that Filipino workers preferred English: because English could be associated with elite social status, some workers experienced its use as “a kind of class-based mockery,” whereas Tagalog was considered “more respectful and more comfortable” (HC01–02). In obstetric and gynecological consultations, a broker reported using Indonesian interpreters who understood workers’ dialects; when the interpreter was female, she could relay sensitive clinical information directly to female workers, reducing misunderstanding (BK01).

Contraceptive acceptability was also shaped by differences between workers’ home-country practices and the options available in Taiwan. An academic participant reported that injectable contraception and implants commonly used by Indonesian women were less available in Taiwan, requiring workers to transition to unfamiliar methods after arrival (AC01). The participant emphasized that workers were not unwilling to use contraception; rather, language, culture, work arrangements, and differences in available methods created a gap between their preferences and the services they could obtain (AC01).

Participants further questioned whether translated SRH education was culturally appropriate. One NGO representative argued that materials needed to reflect “the different cultures and lifestyles of each country,” rather than reproducing the same content in multiple languages (NGO02). Another NGO used concrete, interactive teaching to explain sexual autonomy, gender equality, and legal protections, while staff reported that differing expectations about regular prenatal checkups required additional explanation and reassurance (NGO04–06).

### 3.4 Quality

Quality concerns whether healthcare is clinically appropriate, safely delivered, effectively communicated, and capable of producing continuity of care. [23] Participants generally viewed Taiwan’s clinical services favorably, while identifying weaknesses in interpretation, care coordination, and treatment continuity for migrant workers.

Several participants distinguished these concerns from the technical standard of Taiwan’s healthcare. One healthcare participant stated that migrant workers expressed their “highest satisfaction with Taiwan” in relation to healthcare and sometimes sought to complete treatment before returning home because comparable care might not remain available after return (HC01–02). Another NGO participant similarly described migrant workers’ medical experiences in major cities as “very good” (NGO07).

However, the quality of clinical communication varied substantially. Medical interpretation was described as “extremely lacking,” and participants reported difficulty conveying professional advice and medication instructions accurately across language barriers (AC02; HC03). One healthcare participant questioned whether a migrant worker could “faithfully relay” information between physicians, adding, “I do not know how reliable that is” (HC04). By contrast, one labor-sector participant reported that agency-provided interpreters enabled physicians’ instructions to be communicated clearly and made “the chance of misunderstanding… very small” (BK01). These contrasting accounts indicated that interpretation support materially shaped the reliability and safety of care communication (AC02; BK01; HC03; HC04).

Participants also identified weaknesses in coordination and continuity. Clinical specialties, employment arrangements, insurance issues, and social support were often handled separately, despite requiring an integrated response in complex migrant-worker cases (GOV01; HC04). As one healthcare participant explained, “it is not enough for each specialty simply to handle its own area and be done with it” (HC04). The lack of continuity had an especially negative impact on migrants living with HIV, because delays in obtaining medication could interrupt treatment and worsen outcomes (NGO03). Participants also reported that continuity after return to the country of origin could not be guaranteed, although NGOs sometimes developed individualized care plans or taught workers how to continue treatment and rehabilitation independently (NGO01; NGO02; NGO03).

## 4. Discussions

This study found persistent gaps in migrant workers’ access SRH information, preventive services, and continuity of care despite Taiwan’s universal health insurance and comparatively strong healthcare infrastructure. This pattern is consistent with research on labor migration from low- and middle-income countries (LMICs) to high-income destinations, which has shown that participation in destination-country labor markets does not necessarily ensure equitable access to healthcare. [24–26]

Several findings reflected well-established concerns in this literature. Migrant workers faced difficulties obtaining understandable SRH information, navigating unfamiliar services, and accessing care that corresponded with their linguistic needs and prior health practices. Gaps were particularly evident in contraception, HIV-related care, reproductive health information, and maternal services. The prominent role of NGOs and community organizations in providing education, navigation, interpretation, shelter, and linkage to care also mirrors international evidence that non-state actors frequently compensate for limitations in formal systems serving temporary and mobile labor populations. [27–29]

While these findings align with broader evidence on migrant workers’ health vulnerabilities, our findings also show that these barriers did not occur in isolation. Rather, they reflect interconnected processes through which migrant workers’ SRH needs were translated, negotiated, and sometimes constrained within the destination-country context. We therefore focus our secondary analysis on three dimensions that emerged as central to understanding these processes: how access to SRH care was mediated through labor brokerage; how gendered labor arrangements shaped reproductive autonomy and healthcare experiences among female live-in caregivers; and how fragmented governance shapes the realization of SRH rights within Taiwan’s health system.

### 4.1 Mediation of SRH Care through Brokers

International migration scholarship has long established the mediating role of brokers in migrant workers’ navigation of border control, migration & labor regulations, and life in the destination country. [30–32] Our findings extend this literature into healthcare. In Taiwan, brokers remained important mediators of SRH care even though migrant workers were formally included in NHI system. Read alongside our AAAQ findings, this reliance reflects the various barriers that limited workers’ ability to navigate care independently, including unfamiliarity with the healthcare system, linguistic and cultural differences, and work arrangements that constrained time and mobility.

A government participant described brokers as migrant workers’ first practical point of contact after arrival:

> *“Whenever something happens, the worker turns directly to the broker. As a result, the broker is not only handling employment and documentation. The broker is also expected to serve as interpreter, healthcare navigator, emotional mediator, and sometimes the person who explains medical costs to the employer.” – GOV02*

Brokers’ own accounts corroborated this expanded role. One broker described workers asking the agency for help with initial prenatal visits and pregnancy termination, for which agency staff accompanied them to hospitals for examination and assessment (BK02).

But previous brokerage scholarship also cautions against placing brokers into simple categories of either good or bad actors. [32] In SRH care, brokers can simultaneously facilitate access and create barriers to it. On the one hand, their knowledge, language support, and position within the employment system can help workers reach services they might otherwise struggle to navigate. On the other hand, the involvement of brokers in sensitive SRH care can carry employment and migration consequences. One NGO participant noted that some workers explicitly asked whether their “employer or broker will know” their HIV status (NGO02).

Their concerns were not unfounded. One NGO participant observed that when foreign care workers were found to have HIV, “more often the employer terminates the contract or sends them away,” after which remaining in Taiwan often depended on quickly securing another employer, usually with a broker’s help (NGO03). Despite formal legal protections against HIV-related discrimination, access to remedy remained limited in practice. As one NGO participant explained, workers whose contracts were terminated over their HIV status were often directed by their brokers to find another job or prepare to return home rather than pursuing legal action. “So far we have only encountered one (foreigner) who wanted to do that (use the available remedy process),” said one NGO participant, and that person, as the participant later clarified, was a white-collar foreign researcher (NGO08).

This finding thus makes a distinct contribution to scholarship on the coproduction of precarious migrant labor. Wee et al. conceptualize precarization as the outcome of overlapping formal and informal conditions that migrants must satisfy to maintain security and exercise rights. [30] In Taiwan, the limited availability, accessibility, acceptability, and quality of migrant-responsive SRH services created conditions under which healthcare access could not be realized through formal entitlement alone. Brokers became central because they mediated workers’ encounters with the conditions governing access to SRH care. Under this governance arrangement, migrant workers’ ability to exercise health rights remains contingent on relationships outside the healthcare system.

### 4.2 Precarity of Live-in Domestic Work for Female Migrants

Existing scholarship on female domestic workers paints a richly nuanced picture of live-in caregivers’ SRH health & care-seeking issues (citations). Consistent with this body of literature, our findings show that migrant women face information, linguistic, financial, administrative, and cultural barriers to SRH services.

We argue that all these barriers are shaped by a more fundamental source of precarity: the subordination of migrant women’s reproductive autonomy to the demands of live-in care work. Because workplace and residence coincide, workers’ time, mobility, privacy, and bodily availability are organized around the needs of the care recipient.

One healthcare participant observed:

> *“Because she was a live-in home caregiver, she did not have much time to go out.… Basically, it was a twenty-four-hour arrangement.… If she needed to see a doctor, she had to notify her broker.” – HC01*

Another healthcare participant described the same constraint:

> *“For caregivers it is much harder, because they work one-on-one and have to be beside the patient almost all the time. It is very difficult for them to take leave.… Generally, a migrant caregiver may have only one day off a month.” – HC04*

These accounts show that SRH access is conditioned by more than service availability. The practical ability to attend appointments, obtain follow-up care, or seek confidential advice depends on whether workers can temporarily withdraw from care obligations that are continuous and residential.

The same constraint also shapes intimate and reproductive life outside formal healthcare encounters. One participant described a married caregiver whose husband lived in another city:

> *“They might only see each other once a month, and that one meeting was supposed to be very intimate.” – HC02*

Pregnancy makes this tension especially visible because the worker’s reproductive needs can directly conflict with the physical requirements of care work. As one broker explained:

> *“The person needing care may weigh more than sixty kilograms and need to be lifted.… If she is pregnant, she cannot lift heavy things. So there are only two ways: either the employer keeps her, or the employer does not.” – BK01*

At the extreme, this conflict could extend into reproductive decision-making itself. An academic participant recounted a case in which:

> *“The employer not only disagreed (with the female caregiving taking short leaves to have prenatal checkups), he also requested the migrant worker to go have an abortion, and then requested the broker to take the migrant worker to do it.” – AC02*

Taken together, these findings suggest that the SRH precarity of migrant live-in caregivers arises not from isolated barriers to healthcare, but from an inherent hierarchy embedded in the organization of care work. The live-in arrangement creates an asymmetry in which caregivers are expected to provide continuous attention to the bodily needs of others, while their own bodily needs must be accommodated around the requirements of employment. This asymmetry becomes particularly visible during moments when workers’ bodies no longer align with the expectations of care provision, whether when they require medical attention, maintain intimate relationships, or become pregnant. In these situations, SRH needs are treated as conditions that must be negotiated against the household’s demand for uninterrupted care.

### 4.3 Fragmented & Performative Governance in a High-Performing Health System

Consistent with international literature on migrant health governance, our findings show that NGOs in Taiwan frequently absorb responsibilities created by gaps between SRH policy and implementation, especially when it comes to vulnerable populations. [27–29] Rather than operating only as supplementary service providers, NGOs often became de facto case managers when migrant workers’ needs crossed institutional boundaries. As one participant explained,

> “If the government encounters a foreign person living with HIV and doesn’t know how to handle the situation—especially if the person needs housing—they may come to us. But they cannot provide funding or material support.” –NGO01

These referrals reflected a broader lack of institutional coordination, as the same participant noted that the CDC, immigration authorities, and other agencies often operated independently, leaving no clear operational pathway for complex cases.

Another NGO participant (NGO03) described how this fragmentation played out in practice. In one case, a migrant worker living with HIV was required to remain in Taiwan during legal proceedings, yet immigration authorities could not place her in existing shelters and no other agency assumed responsibility. The NGO ultimately provided housing, food, and limited living support through its halfway house (a program created for migrants in between jobs or legal status). As the participant summarized, “when migrant workers face these sorts of situations, there is often no clarity about who is supposed to take care of them.”

In fact, formal coordination mechanisms did exist, but participants suggested that their effects were often more procedural than substantive. Several NGO participants reported their participation in the Ministry of Health and Welfare Committee on HIV/AIDS Prevention and the Protection of the Rights of People Living with HIV, which brings government agencies together with civil-society organizations to discuss HIV prevention and rights protection. NGO03 described its role as bringing frontline experience into this process: “Our role there is to respond to the Ministry of the Interior by saying, based on what we see in our field of service, what is lacking and what needs to be strengthened.” The committee could generate concrete administrative responses; NGO03 noted that when members raised concerns, the Ministry of the Interior would subsequently report what revisions it had made.

However, NGO08’s account suggests that this coordination could become highly performative. For an issue involving misinformation about HIV and migrant-worker recruitment, the participant repeatedly raised evidence with the Ministry of Labor, yet reported having to “keep pushing, bit by bit,” with responses that did not necessarily address the population affected. The participant described the committee process itself as dependent on formal tracking: “the first step is getting the proposal into the meeting minutes. Only then can it become a tracked case… If it is not tracked, it is as if it does not exist.”

Even when issues were formally assigned for follow-up, substantive change was not guaranteed. After the NGO proposed HIV-specific in-service training for foreign care workers, “It did not happen”; they did, nevertheless, get a response saying “HIV was already covered under broader blood-borne disease training and that should be enough.” –NGO08

### 4.4 Policy Recommendations

We recommend reforms that strengthen both migrant workers’ formal SRH protections and the institutional mechanisms needed to exercise them. First, Taiwan should reduce dependence on brokers for healthcare navigation by creating an independent migrant-health navigation function accessible through hospitals, local health departments, and the 1955 hotline. This should include trained multilingual interpreters, confidential SRH counseling, and referral support that workers can access without employer or broker involvement. Hospitals treating migrant workers for HIV, pregnancy, or other sensitive SRH conditions should be able to refer directly to this service rather than relying primarily on brokers or informal NGO networks.

Second, we recommend strengthening the implementation of existing protections for live-in caregivers. Taiwan already prohibits pregnancy-based dismissal and provides formal complaint and assistance channels, but these protections are harder to exercise when workers live inside the employer’s household and depend on employers or brokers for mobility, information, and continued employment. Enforcement should therefore focus on making existing protections independently accessible. This could include routine multilingual information on pregnancy and sexual-harassment rights, direct referral from hospitals and the 1955 hotline to labor authorities, and rapid access to temporary accommodation or employer transfer when remaining in the household becomes unsafe or untenable.

Third, we recommend strengthening integrative governance capacity across the institutions that shape migrant SRH access. Building on Taiwan’s existing healthcare infrastructure—along with its specialized agencies, referral mechanisms, and interagency committees— policymakers should establish clearer cross-agency case-management pathways. At the policy level, interagency coordination should also be evaluated by implementation outcomes, not merely by whether issues are recorded, assigned, or formally answered.

### 4.5 Limitations

While our research provides valuable insights into the realization of migrant workers’ right to sexual and reproductive health in Taiwan, it is not without limitations. First, our relatively small purposive sample included only a limited number of representatives from each stakeholder group. The findings therefore capture a range of institutional perspectives but cannot represent the full diversity of experiences across Taiwan’s healthcare, labor, brokerage, government, and civil-society sectors. Second, migrant workers’ own perspectives were not directly included because of the study’s IRB-approved recruitment scope. Our analysis consequently reflects how intermediaries and institutional actors perceived migrant workers’ needs and barriers, rather than workers’ own accounts of care-seeking, autonomy, and decision-making. Third, the thematic range of the data was uneven. HIV and pregnancy-related issues were discussed most extensively, whereas sexual harassment, sexually transmitted infections, intimate relationships, and broader sexual-health experiences appeared less frequently. This may partly reflect the sensitivity and privacy of these issues, which can make them less visible to the intermediaries and service providers interviewed. Future research should therefore include migrant workers directly and examine a broader range of SRH experiences across worker groups and employment settings.

## 5. Conclusion

In conclusion, this study highlights persistent gaps in the realization of migrant workers’ right to sexual and reproductive health in Taiwan despite broad health coverage and comparatively strong healthcare infrastructure. Addressing these gaps requires attention to all four dimensions of the AAAQ framework: availability, accessibility, acceptability, and quality. In particular, migrant workers need more direct access to understandable information, confidential and culturally responsive services, and continuity of care that does not depend excessively on employers, brokers, or NGOs. For female live-in caregivers, stronger implementation of existing protections is needed to ensure that reproductive health needs, pregnancy, and sexual harassment concerns can be addressed without jeopardizing employment or residence. Finally, more coherent cross-agency governance is essential so that formal rights are matched by clear responsibility and effective follow-through. A rights-based approach to migrant SRH therefore requires both robust legal protections and institutional arrangements capable of translating those protections into practice.

## Data Availability

All data produced in the present work are contained in the manuscript

## Ethics Declaration

Ethics approval and consent to participate: This study was conducted in accordance with the ethical principles of the Declaration of Helsinki. Ethical approval was obtained from the Harvard Longwood Campus Institutional Review Board (IRB25-1054) under exempt review. All participants provided verbal informed consent prior to participation.

## Funding Declaration

This study was supported by grant funding from the Population Health and Welfare Research Center at National Taiwan University. The funder had no role in the study design, data collection, analysis, interpretation of the findings, or preparation of the manuscript.

## Notes

### Competing Interest Statement

The authors have declared no competing interest.

### Author Declarations

The Institutional Review Board at Harvard Longwood Campus (IRB25-1054) gave ethical approval for this work

